# Improving patient clustering by incorporating structured label relationships in similarity measures

**DOI:** 10.1101/2023.06.06.23291031

**Authors:** Judith Lambert, Anne-Louise Leutenegger, Anaïs Baudot, Anne-Sophie Jannot

**Author notes:** Corresponding author: Judith Lambert, PariSanté Campus, 10 rue d’Oradour-sur-Glane, 75015 Paris, France. These authors contributed equally to this work (Anaïs Baudot, Anne-Sophie Jannot).

## Abstract

**Context:** Patient stratification is the cornerstone of numerous health studies, serving to enhance medicine efficacy estimation and facilitate patient matching. To stratify patients, similarity measured between patients can be computed from medical health records databases, such as medico-administrative databases. Importantly, the variables included in medico-administrative databases can be associated with labels, which can be organized in ontologies or other classification systems. However, to the best of our knowledge, the relevance of considering such label classification in the computation of patient similarity measures has been poorly studied.

**Objective:** We propose and evaluate several weighted versions of the Cosine similarity that consider structured label relationships to compute patient similarities from a medico-administrative database.

**Material and Methods:** As a use case, we analyze medicine reimbursements contained in the *Échantillon Généraliste des Bénéficiaires*, a French medico-administrative database. We compute the standard Cosine similarity between patients based on their medicine reimbursement. In addition, we computed a weighted Cosine similarity measure that includes variable frequencies and two weighted Cosine similarity measures that consider label relationships. We construct patient networks from each similarity measure and identify clusters of patients. We evaluate the performance of the different similarity measures with enrichment tests using information on chronic diseases.

**Results:** The similarity measures that include label relationships perform better to identify similar patients. Indeed, using these weighted measures, we identify distinct patient clusters with a higher number of chronic disease enrichments as compared to the other measures. Importantly, the enrichment tests provide clinically interpretable insights into these patient clusters.

**Conclusion:** Considering label relationships when computing patient similarities improves stratification of patients regarding their health status.

## 1 INTRODUCTION

Identifying similar patients can serve multiple purposes in healthcare. In routine practice, finding patients similar to a given patient is often beneficial, particularly in the case of rare diseases (1). Similar patients may share the same condition, and can provide diagnostic and prognostic guidance. In research, identifying groups of similar patients is useful to stratify the population. This enables, for instance, a more precise estimation of medicine efficacy for a given patient profile or matching similar patients in case-control studies (2,3).

The similarity between patients can be computed from medical health records such as medico-administrative databases. Medico-administrative databases contain data used for health care reimbursement purposes, including information about hospitalization, medicine and medical device consumption. They therefore provide a comprehensive perspective on the entire healthcare pathway of a given patient.

The variables contained in these databases are labeled by terms that can be related to each other. For instance, medicines are labeled by their names and organized into classifications such as the Anatomical Therapeutic Chemical (ATC) classification. Within this classification, all anti-diabetic medicines belong to the same class. Thus, the labels of two medicines used to treat diabetes are related according to the classification, and patients treated with these medicines are expected to be more similar than patients treated with medicines from different classes. Other classification systems are available for various types of medical data. For instance, the International Classification of Diseases (ICD) is used for diagnoses (4) and SNOMED-CT is used for clinical information (5).

The objective of similarity measures is to identify patients sharing characteristics, such as taking the same medicine or having the same diagnoses at a specific age. The most commonly used measures to compute similarities between patients are the Euclidean distance, the Jaccard index, and the Cosine similarity (6). Weighted measures can also be incorporated to these similarity measures to account for the frequency of variables. For instance, the Inverse Document Frequency is a weighted measure that assigns greater importance to rare variables (7,8). However, these classical similarity measures rely only on the variable values and do not consider other information associated with the variables, such as their labels. For instance, when considering medications, two patients are similar if they take similar dosage of similar medicine. In this case, similarity is defined at both the dosage level (i.e., variable value) and at the medicine level (i.e., variable label relationships). Hence, analyzing the relationships between variable labels, by leveraging a label classification system, is expected to provide pertinent information for computing patient similarity from a clinical perspective.

Several measures have been proposed to analyze label relationships. The Wu and Palmer measure examines the relationships between two labels by considering their depth in the classification (9), while the Lin measure considers both their depth and their frequency in the classification (10). These measures have been incorporated into the computation of patient similarities in various studies. For instance, Ni et al. compute patient similarities based on their ICD-10 diagnosis, using a weighted similarity measure that considers the classification depth of ICD-10 codes (11). Girardi et al. adapted the Jaccard distance to include diagnose relationships in patient similarity computation based on the depth of their ICD-10 codes (12). However, these weighted patient similarity measures are limited to variables associated with binary values (i.e., Boolean variables) and cannot be applied to quantitative variables. To the best of our knowledge, similarity measures able to simultaneously consider label relationships from classifications and quantitative values of variables are lacking in the field.

The efficiencies of the similarity measures are usually estimated by assessing the quality of the clusters obtained using the measures. The clustering performance is evaluated with metrics such as silhouette score or accuracy. However, interpreting these performance metrics from a clinical perspective can be challenging. An alternative method to assess the performance of clustering involves using external variables that were not used initially to compute patient similarities. These external variables can be related to prognosis (13) or tumor characteristics (14), for instance.

In this paper, we propose to weight the Cosine similarity to include label relationships in order to identify similar patients. We further aim to assess the added value of incorporating this information thanks to an evaluation protocol that can be interpreted clinically. Our study focuses on a specific use case related to medicine consumption in a national French medico-administrative database. We first compute several weighted similarity measures and employ them to cluster patients, thereby revealing groups of similar patients. We then assess the performance of the different similarity measures in identifying clusters of patients thanks to enrichment tests based on external variables (i.e., diagnoses). We observed that taking into account the relationships between the labels in the computation of patient similarities improves the quality of the identified patient clusters.

## 2 MATERIAL AND METHODS

Let I be the set of variables (i.e., medicines). Let *X* and *Y*, the vectors of variables from *I* for two patients. We compute the similarity between patients using four different measures. The first two measures (Cosine similarity and Cosine similarity weighted by the Inverse Document Frequency) rely on quantitative variables, while the remaining two (Cosine similarity weighted by the Wu and Palmer measure and Cosine similarity weighted by the Lin measure) rely on both quantitative variables and label relationships of the variables.

### 2.1 Defining patient similarity measures

#### 2.1.1 Cosine similarity

The Cosine similarity between two patient vectors *X* and *Y* is defined as the cosine of the angle (*θ*) between the two vectors (15):

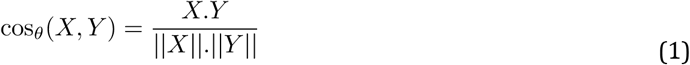

The Cosine similarity values range from -1 to 1, with value equal to -1 when vectors are opposite, 0 when vectors are different (i.e., orthogonal) and 1 when they are identical.

#### 2.1.2 Cosine similarity weighted by the Inverse Document Frequency

The Cosine similarity weighted by the Inverse Document Frequency (IDF) is defined as follows for two patient vectors *X* and *Y* (7):

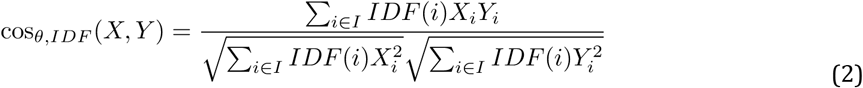

with 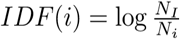, where *N*_*I*_ is the total number of observations of all the variables in the set *I* and *N*_*i*_ is the total number of observations of the variable *i*.

As for the standard Cosine similarity, the values of this weighted version range from -1 to 1.

#### 2.1.3 Cosine similarity weighted by the Wu and Palmer measure

The Cosine similarity weighted by the Wu and Palmer measure is defined as follows for two patient vectors *X* and *Y* (9):

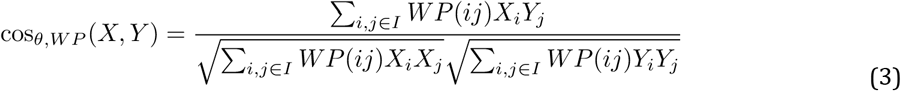

with *WP*(*i,j*) being the Wu and Palmer measure between the labels of the two variables *i* and *j* of the set *I*.

The labels of the variables of the set *I* are organized into a classification consisting of successive levels (*Figure 1*). The label composing the top level is the root and the labels composing the lowest level are the leaves. Each level in the classification is connected to the next level through edges, representing the relationship between labels in the classification. A sequence of edges represents a path in the classification.

**Figure 1:**
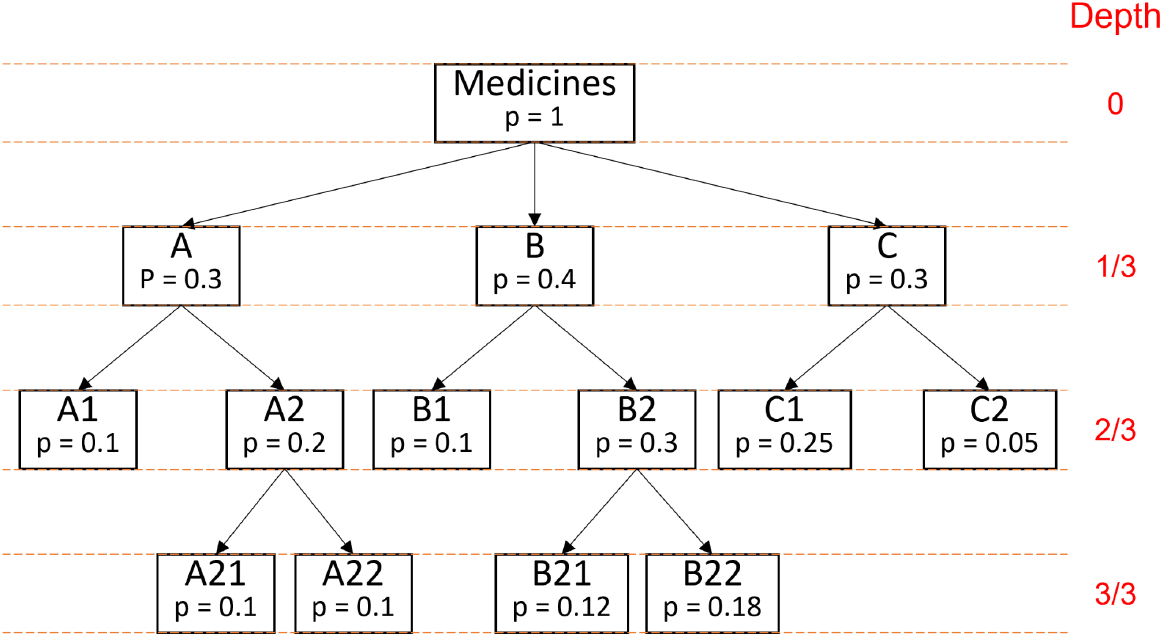
Example of a classification of medicine data The classification is composed of several medicine labels organized in successive levels interconnected by edges. The depth of a given variable label is the number of edges between the root (i.e., label “Drugs”) and that given variable label, divided by the number of edges in the shortest path from the root to the leaves. *p* is the medicine label frequency

The Wu and Palmer measure is computed from the variable labels as follows:

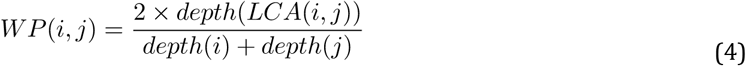

with *depth*(*z*) = *E*_*z*_*/E* where *E*_*z*_ is the number of edges between the root and the variable label *z* in the classification and *E* is the total depth in the classification (i.e., the number of edges in the shortest path from the root to the leaves); *LCA*(*i,j*) is the Lowest Common Ancestor of the labels of the variables *i* and *j* in the classification (i.e., the lowest label of the variable of set *I* that has both *i* and *j* as descendants). For example, the Wu and Palmer measure between the medicine labels B1 and B22 from the classification of the *Figure 1* is computed as follows:

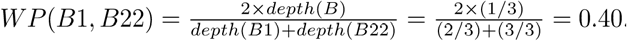

As for the standard Cosine similarity, the values of this weighted version range from -1 to 1.

#### 2.1.4 Cosine similarity weighted by the Lin measure

The Cosine similarity weighted by the Lin measure is defined as follows for two patient vectors *X* and *Y*:

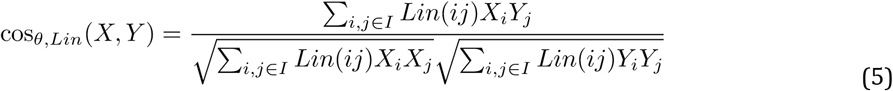

With *Lin*(*i,j*) the Lin measure between the labels of the two variables *i* and *j* of the set *I*.

The Lin measure analyzes the relationship between two variables *i* and *j* by considering the information content (IC) of the labels of the two variables and the information content of their lowest common ancestor (10):

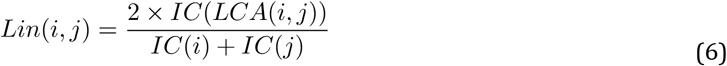

with *IC*(*z*) = −log*P*(*z*) where *P*(*z*) is the probability of occurrence of the variable *z* estimated by its frequency. For example, the Lin measure between the medicine labels B1 and B22 from the classification of the *Figure 1* is computed as follows:

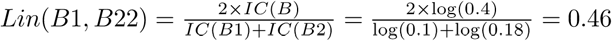

As for the standard Cosine similarity, the values of this weighted version range from -1 to 1.

### 2.2 Identifying clusters of patients from patient networks

Various clustering methods can be used to identify clusters of patients from their similarity measures. Some examples include Kmeans, hierarchical clustering, or the Markov cluster algorithm applied to patient networks (16,17). In a previous work, we showed that building patient networks using Cosine similarity on medicine data and clustering the networks was a pertinent approach to identify patient clusters and trajectories (18). Therefore, here, we build patient networks using Cosine similarity or its weighted versions on medicine data, and cluster the networks with the Markov Cluster algorithm to identify clusters of patients.

#### 2.2.1 Constructing patient networks

A patient network is a graph *G* = (*V,E*) with *V* patient nodes and *E* edges representing interactions between patient nodes. The network is constructed using a similarity matrix. Let *M* = [*m*_*X,Y*_]^*n*^ be the similarity matrix where *n* is the number of patients and *m*_*X,Y*_ is the similarity between patient vectors *X* and *Y*. This similarity matrix is symmetrical, with *m*_*X,Y*_ = *m*_*Y,X*_. We compute four similarity matrices, each corresponding to a specific similarity measure. We then apply a threshold *t* to the similarity matrices to construct the patient networks. Two patients are connected in the network (i.e., an edge between the patients is present) if their similarity is above the threshold *t*. The connection between patients X and Y is weighted by the value *m*_*X,Y*_ of the matrix.

To ensure comparable networks for the different similarity measures, we select a distinct threshold *t* for each measure. These thresholds are chosen to obtain approximately 5000 patient nodes in the largest connected component of each network (*supplementary Table S1*).

#### 2.2.2 Clustering patient networks

We apply the Markov Cluster algorithm (MCL) (19) on the largest connected component of each patient network. The MCL algorithm uses random walks to simulate flows on the network. The flows allow to distinguish network areas where nodes are strongly connected, which correspond to the clusters. We use the version 0.0.6.dev0 of the “markov-clustering” Python package with the default parameters.

### 2.3 Cluster enrichment analysis

Let external variables be binary variables that are not used to compute the similarities between patients. The aim of the enrichment analysis is to assess if each external variable has a frequency higher than expected in a cluster. For each external variable and each cluster of patients, we compare patients inside and outside the cluster using Fisher’s exact test (20). This procedure involves performing a number of tests equal to the product of the number of clusters times the number of external variables. We adjust this multiple testing with the Benjamini-Hochberg procedure. We consider that a variable is enriched in a given cluster if its adjusted p-value is lower than 0.05.

### 2.4 Use-case: the Échantillon Généraliste des Bénéficiaires

We use health data from the Échantillon Généraliste des Bénéficiaires (EGB), a French medico-administrative database. The EGB is a random sample of the French health insurance database (21). It is representative of the French population and contains approximately 660,000 individuals followed over a period of 11 years. According to French regulation, individuals in SNDS (Système National des Données de Santé) database are informed of the reuse of their data for research and can oppose this reuse as defined by Articles 92 to 95 of Decree No. 2005-1309 of 20 October 2005 (https://www.legifrance.gouv.fr/loda/article_lc/LEGIARTI000037300884/). As required by the French regulation, EGB data can be reused for research projects from authorized persons once the research project is declared to their institution (in our case, INSERM).

We extract from the EGB data on medicine reimbursements between 2008 and 2018 (*Figure 2*), including the date of reimbursement and the medicine classification in the Anatomical Therapeutic Chemical (ATC) class (see example *Table 1*). The ATC class is an international classification of medicines established by the World Health Organization (WHO) (22). We exploit this classification in the patient similarity measures. We then select patients aged 60 and who had received reimbursement for at least one medicine for two or more consecutive months. We therefore keep only patients with sustained reimbursements. We also extract diagnostic data about chronic diseases declared by the patient to the French health insurance. Diagnoses are coded with the 10*th* revision of the international statistical classification of diseases and related health problems (i.e., using ICD-10 code). We thus exclude from our analysis the patients with no declared chronic diseases. Notably, diabetes emerges as the most frequent chronic disease observed within the population. We analyze female and male patient datasets separately. In each dataset, we calculate for each patient, the number of reimbursements they had for each medicine at age 60 (see example *Table 2*).

**Table 1:**
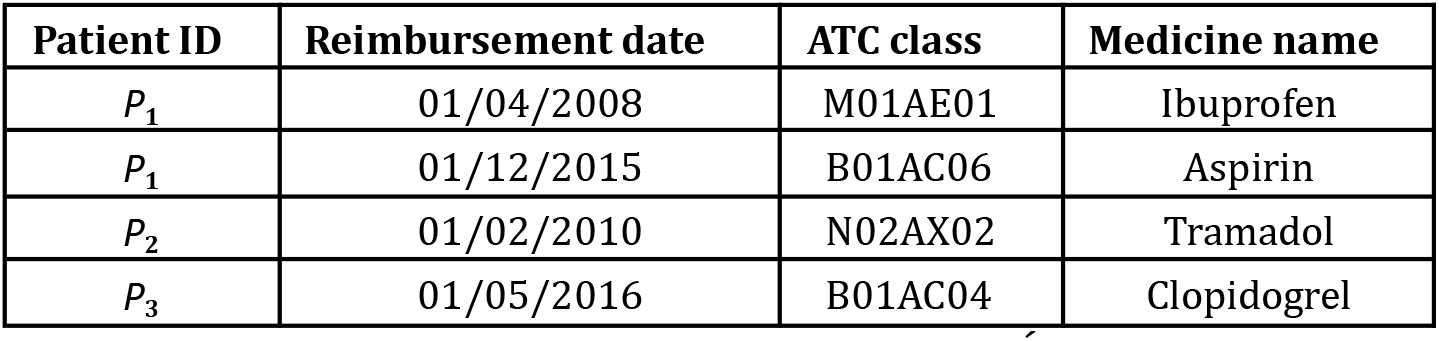
Example of medicine reimbursements contained in the Échantillon Généraliste des Bénéficiaires (EGB) ATC: Anatomical Therapeutic Chemical

**Table 2:** Example of total number of reimbursements that three patients aged 60 years received for three different medicines

| Patient ID | Tramadol | Aspirin | Ibuprofen |
| --- | --- | --- | --- |
| $P_1$ | 0 | 10 | 5 |
| $P_2$ | 1 | 8 | 4 |
| $P_3$ | 2 | 6 | 3 |

**Figure 2:**
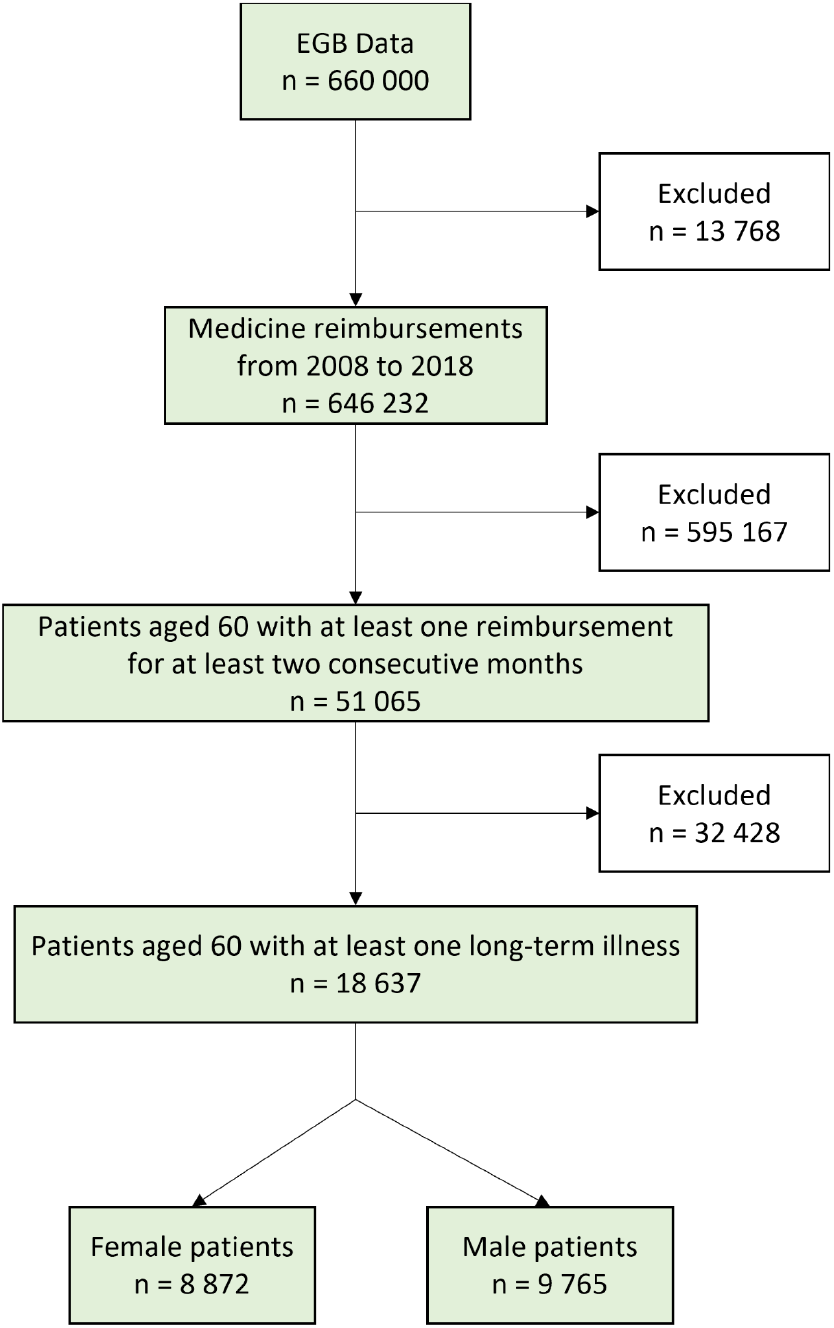
Flowchart of the medicine data extraction process from the Échantillon Généraliste des Bénéficiaires (EGB)

## 3 RESULTS

Our two use-case datasets are composed of 8,872 female and 9,765 male patients. For each dataset, we compute the similarity between patients, build networks, and identify clusters. We assess the performance of the different patient similarity measures with enrichment tests on the patient clusters using declared chronic diseases.

### 3.1 Similarity measures including label relationships have higher similarity values in the use-case populations

We first compare patient similarities computed from medicine reimbursements using four similarity measures, i.e., the standard Cosine similarity and its weighted versions (*Material and methods 2*.*1*).

In the dataset of female patients, the Cosine similarity weighted by the Wu and Palmer measure and the Cosine similarity weighted by the Lin measure identify more patient pairs with similarities with non-zero values (*n*_*0*_ = 3.89×10^7^ for these two measures) as compared to the Cosine similarity and the Cosine similarity weighted by IDF (*n*_*0*_ = 3.02×10^7^ for the two other measures) (*Figure 3*). Additionally, these two weighted Cosine similarity show a higher variability. Thus, the Cosine similarity weighted by the Wu and Palmer measure and the Cosine similarity weighted by the Lin measure, which both include label relationships information, have higher similarity values. Similar results are observed in the dataset of male patients (*Figure S1*).

**Figure 3:**
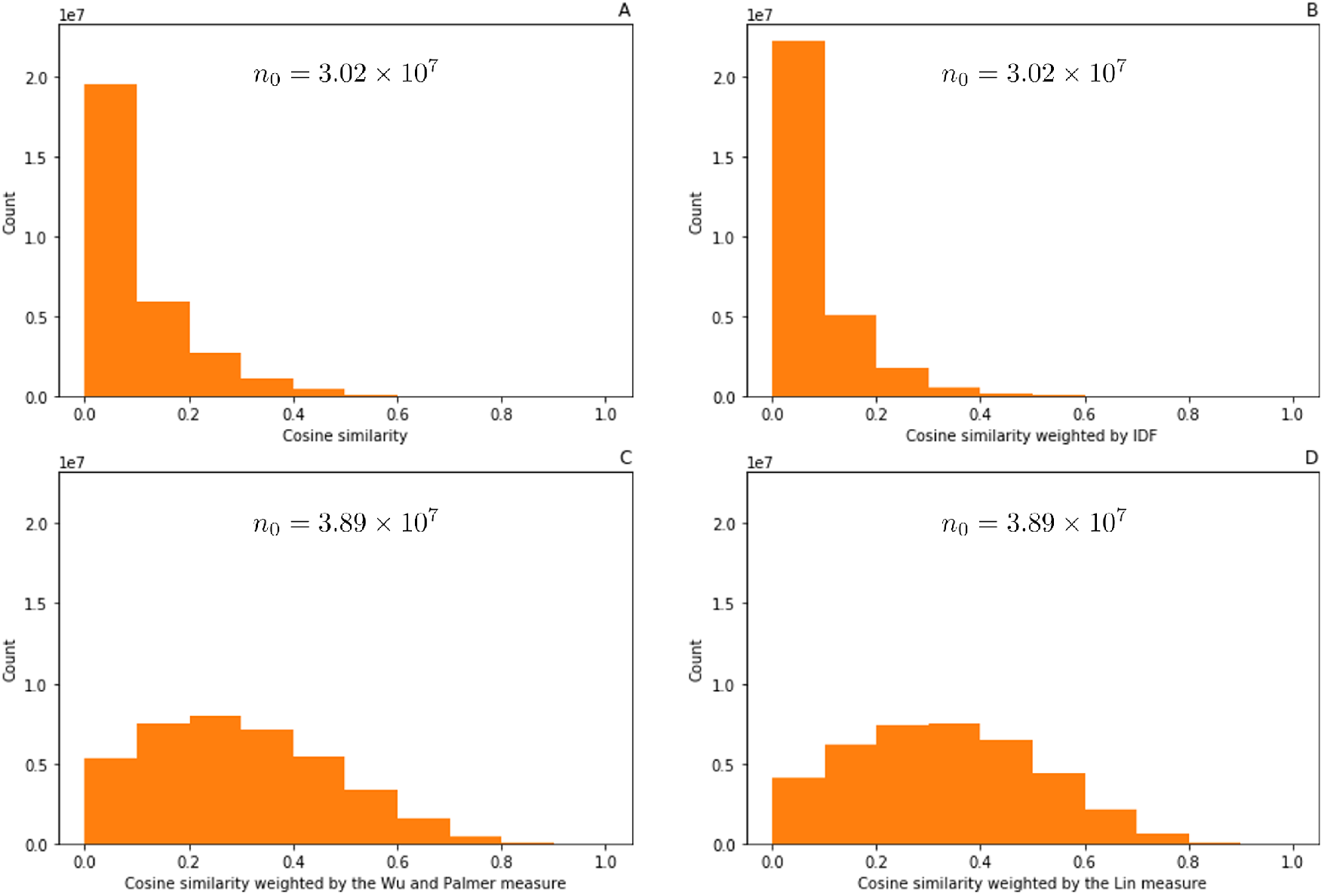
Similarity distributions in the female patient dataset A: Distribution of the Cosine similarity, B: Distribution of the Cosine similarity weighted by the Inverse Document Frequency (IDF), C: Distribution of the Cosine similarity weighted by the Wu and Palmer measure, D: Distribution of the Cosine similarity weighted by the Lin measure. *n*_0_: Total number of patient pairwise similarities with non-zero values.

### 3.2 Similarity measures including label relationships improve patient cluster quality

A patient network is constructed for each of the four similarity measures, in both male and female datasets, leading to 8 different patient networks (*Material and methods 2*.*1*). The *Figure 4* shows the two networks constructed for the dataset of female patients using the Cosine similarity and the Cosine similarity weighted by the Wu and Palmer measure. The network constructed with the Cosine similarity (*Figure 4*A) displays a highly connected structure. Conversely, the network constructed with the Cosine similarity weighted by the Wu and Palmer measure (*Figure 4*B) reveals distinct subnetworks.

**Figure 4:**
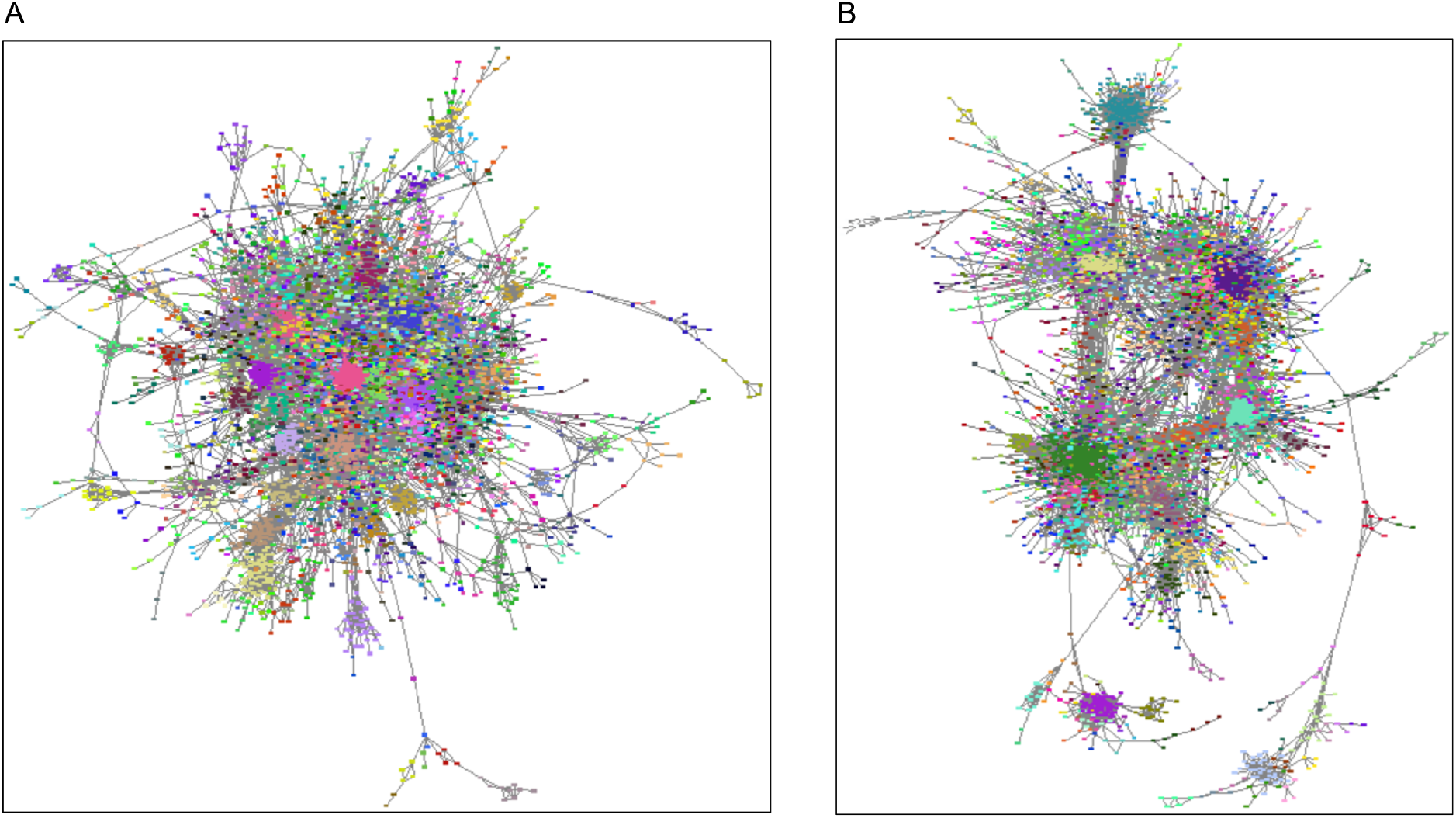
Patient networks built from Cosine similarity and Cosine similarity weighted by the Wu and Palmer measure Networks are built from Cosine similarity (A) and Cosine similarity weighted by the Wu and Palmer measure (B), on the female patient dataset. Nodes represent patients aged 60 and edges represent the interactions between those patients. The length of edges is inversely proportional to the Cosine similarity or the Cosine similarity weighted by the Wu and Palmer measure. Node colors represent the clusters identified with the Markov Clustering algorithm. For the sake of visualization, we only represent the largest connected component of each network.

In the network of female patients built with the Cosine similarity, we identify 12 clusters composed of at least 50 patients. We carry out an enrichment analysis to identify, in each cluster, potential enrichments in chronic diseases (Figure 5A). The enrichment analysis reveals several significant enrichments. For instance, we observe significant enrichments of patients with thyroid and breast cancers in cluster 1, cerebrovascular diseases in cluster 5 and depressive episodes in cluster 2. Similarly, in the dataset of male patients, we identify 11 clusters (*Figures 6A*). The enrichment analysis reveals significant enrichments of patients with cerebrovascular diseases in cluster 5, atherosclerosis in cluster 6, prostate cancer in cluster 8 and thyroid cancer in cluster 9. Of note, we identify several clusters with the same enriched chronic diseases. For instance, female clusters 2, 3, 9 and 11, and male clusters 2, 4, 7, 10 and 11 are all enriched in type 2 diabetes patients. Female clusters 6 and 7 are enriched in breast cancer patients, female clusters 10 and 12 in autoimmune disorder patients and male clusters 1 and 3 in coronary diseases patients. Overall, the use of Cosine similarity allows to identify clusters of similar patients. However, several clusters are redundant regarding their chronic disease enrichments. Similar results are obtained with the Cosine similarity weighted by IDF (*Figures 5 B and 6 B*).

**Figure 5:**
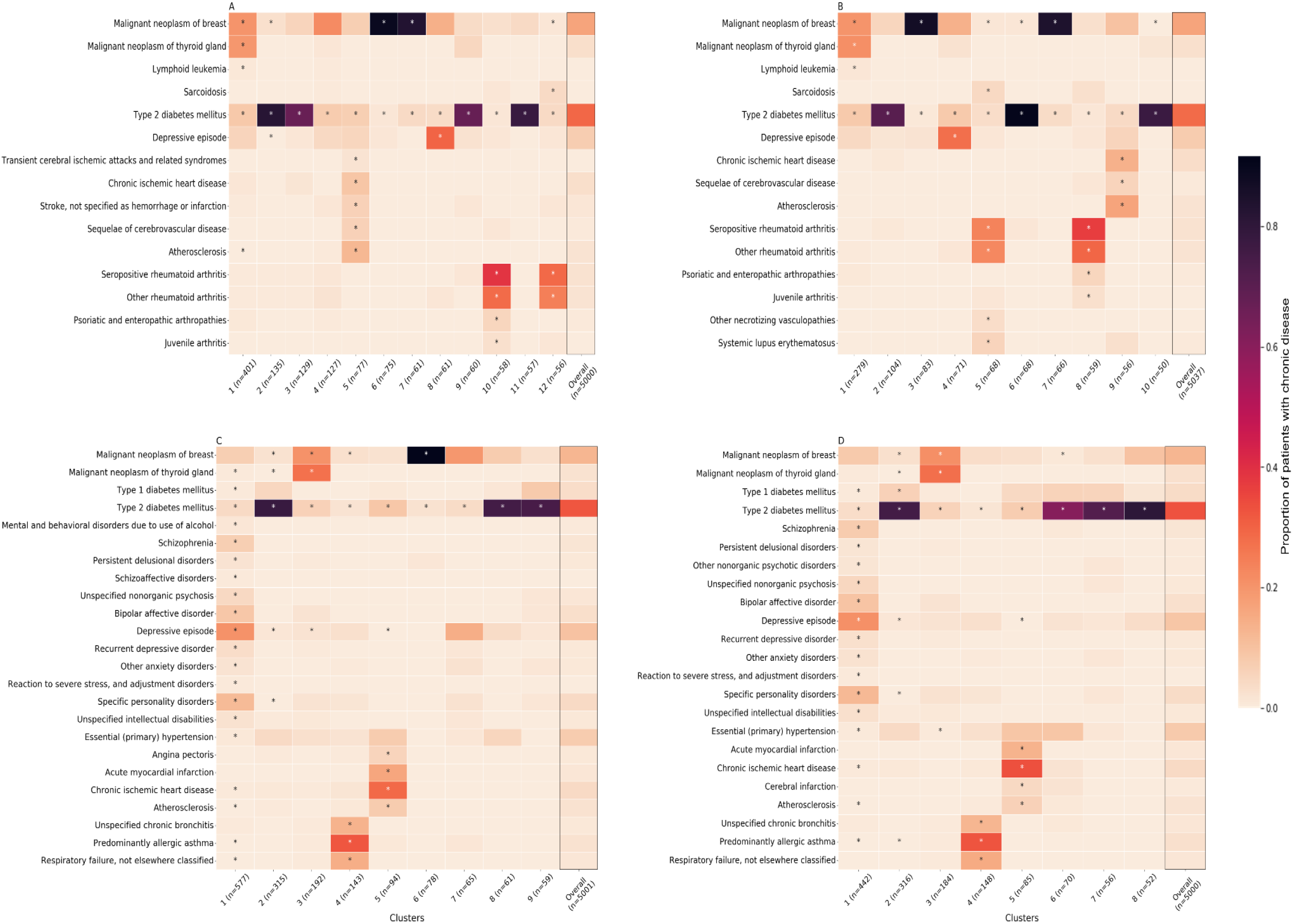
Chronic disease enrichments in patient clusters obtained from the female patient dataset Clusters are identified in networks built from Cosine similarity (A), Cosine similarity weighted by the Inverse Document Frequency (B), Cosine similarity weighted by the Wu and Palmer measure (C), and Cosine similarity weighted by the Lin measure (D), on the female patient dataset. The numbered columns represent the clusters composed of at least 50 patients, ranked from the largest to the smallest. The last column, named overall, represents all the patients found in the network largest connected component. We observe that diabetes is the most frequent chronic disease. n: number of patients identified in each cluster or in the network largest connected component. The rows correspond to the chronic diseases. Box colors represent the proportion of patients with a given chronic disease. Stars represent significant enrichments (p-value lower than 0.05 after Benjamini-Hochberg correction). For the sake of visualization, we only represent chronic diseases that are significant in at least one cluster.

**Figure 6:**
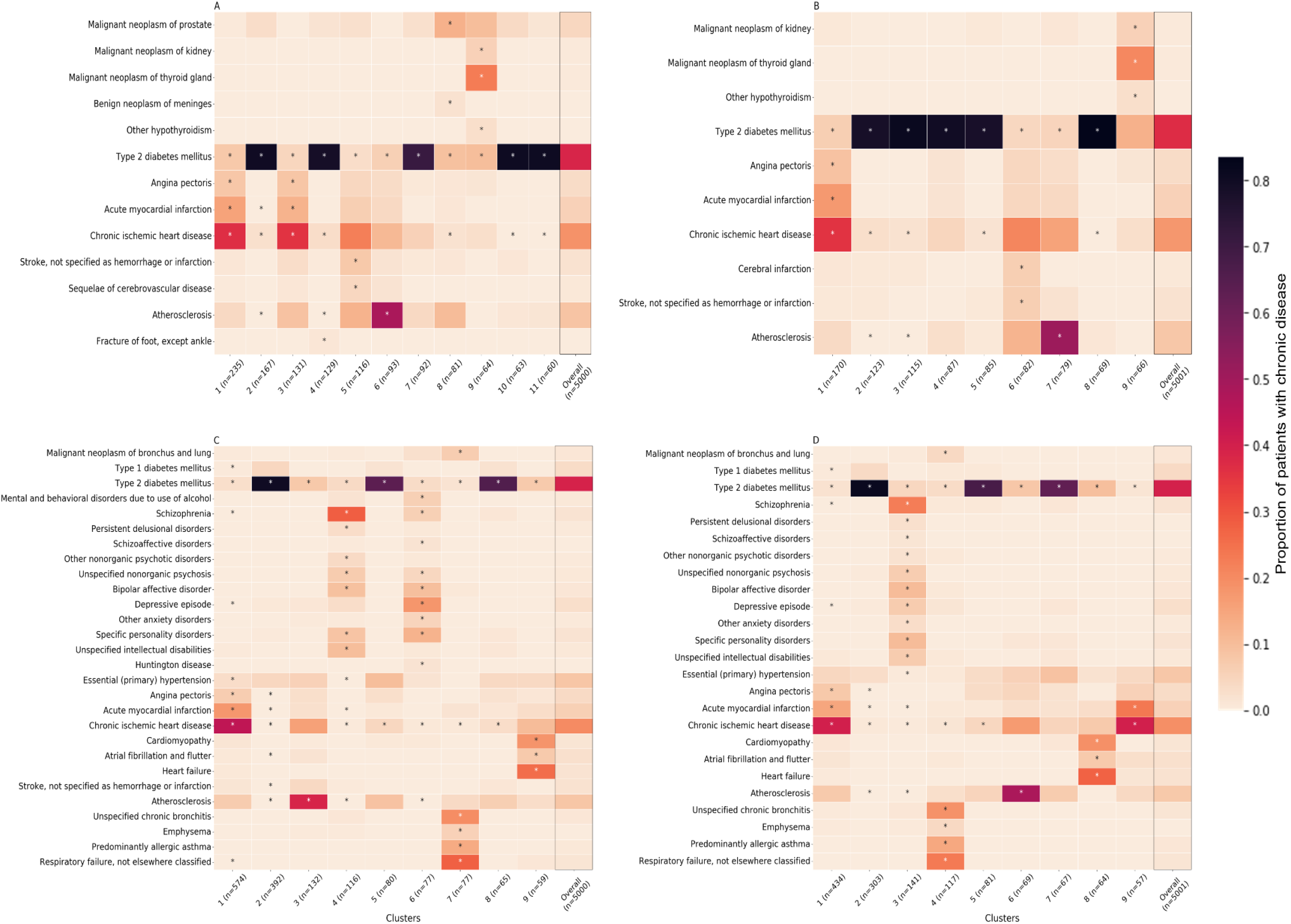
Chronic diseases enrichments in patient clusters obtained from the male patient dataset Clusters are identified in networks built from Cosine similarity (A), Cosine similarity weighted by the Inverse Document Frequency (B), Cosine similarity weighted by the Wu and Palmer measure (C), and Cosine similarity weighted by the Lin measure (D), on the male patient dataset. The numbered columns represent the clusters composed of at least 50 patients, ranked from the largest to the smallest. The last column, named overall, represents all the patients found in the network largest connected component. We observe that diabetes is the most frequent chronic disease. n: number of patients identified in each cluster or in the network largest connected component. The rows correspond to the chronic diseases. Box colors represent the proportion of patients with a given chronic disease. Stars represent significant enrichments (p-value lower than 0.05 after Benjamini-Hochberg correction). For the sake of visualization, we only represent chronic diseases that are significant in at least one cluster.

Patient networks constructed with the Cosine similarity weighted by the Wu and Palmer measure and the Lin measure result in a higher number of clusters significantly enriched in chronic diseases and less redundant clusters (*Figures 5C, 5D, 6C and 6D*). Using the Cosine similarity weighted by the Wu and Palmer measure (*Figures 5C for the female dataset and 6C for the male dataset*), the enrichment analysis reveals clusters significantly enriched in respiratory disease patients (female cluster 4 and male cluster 7), psychiatric disorders patients with psychotic side (female cluster 1 and male cluster 6), coronary disease patients (female cluster 5 and male cluster 1) and patients with type 2 diabetes associated with its comorbidities (female and male clusters 2). In the dataset of female patients, we find clusters significantly enriched in thyroid and breast cancer patients (clusters 3 and 6). In the dataset of male patients, we find clusters significantly enriched in atherosclerosis patients (cluster 3), depressive disorders patients (cluster 4) and heart failure patients (cluster 9). Similar results are obtained using the Cosine similarity weighted by the Lin measure (*Figures 5D and 6D*).

Notably, all clusters are significantly enriched in diabetes patients in both datasets. This is explained by the fact that diabetes is the most frequent chronic disease in our population (see overall columns in *Figures 5 and 6*)

## 4 DISCUSSION

In this paper, we proposed two novel weighted similarity measures for health quantitative variables. These similarity measures were weighted by considering the variables’ labels relationships. They performed better in identifying clusters of patients suffering from different diseases as compared to unweighted similarity measures that did not consider label relationships. Overall, our analysis highlighted the interest of considering label relationships when calculating patient similarities to improve patient stratification.

In recent years, there has been a growing interest in computing patient similarities using Electronic Health Records (EHR). However, most papers focused on computing similarities using variables extracted from medical texts through natural language processing methods. These papers also focused on the development of methods to automatically learn label relationships from data (23). However, using label relationships from existing medical classifications has been poorly addressed, despite the ready availability of this expert information. In this study, we underline the value of integrating such an expert knowledge into patient similarity measures to increase clustering performance. This is particularly relevant to analyze records obtained from administrative claim databases. Indeed, these databases gather medical variables with labels that are always organized into classifications such as SNOMED-CT or ICD-10.

While our study only considered label relationships organized according to a classification, other types of label organization exist. For instance, the Human Phenotype Ontology (HPO) is a directed acyclic graph (DAG). Previous studies have already proposed label relationship measures for this type of label organization. For example, Köhler et al. developed a label relationship measure that exploits the structure of HPO to improve clinical diagnostics (24). Xue, Peng, and Shang derived another measure that exploits both the DAG structure and the phenotype term definition of HPO in order to improve the prediction of disease-related phenotypes (25). However, these measures were originally designed for binary variables and would need to be adapted for quantitative variables commonly found in medical health records as well as in many biological and omics datasets. A recent work demonstrated the interest of considering prior knowledge representation in the context of omics data (13).

In this study, we used the Cosine similarity because we have previously shown that this measure was more effective than others to deal with our specific use-cases (18). However, depending on the data, other similarity measures could be used, and weighted, to compute similarities between patients. We also explored the interest of incorporating variable frequencies in the computation of patient similarities. Indeed, we weighted the Cosine similarity by the Inverse Document Frequency (IDF) to take into account the frequency of the usage of the medicines. Our hypothesis was that it would better capture similarity information as two patients taking the same uncommon medicine would be considered more similar than two patients taking the same common medicine. However, we observed that exploiting the medicine frequency did not enhance the performance of the similarity measures. This may be attributed to the fact that a single medicine can be used to treat multiple conditions, making it challenging to associate a medicine with a specific pathology. However, considering the frequency of medicines may be more effective in the context of rare diseases (26). In such cases, these diseases are typically treated with orphan medicines that have specific indications.

In this paper, we assessed the performance of the different similarity measures to cluster patients using external binary variables (i.e., chronic diseases). We employed these external variables in cluster enrichment analyses. This novel approach deviates from the typical reliance on internal criteria such as silhouette score, which do not offer clinically interpretable insights. Using these enrichment analyses, we were able to interpret the clusters clinically and to compare the different similarity measures from an expert point of view. Although previous works have already used enrichment analyses to study enrichments of HPO terms in the literature (27), to the best of our knowledge, it was never used on patient medical data.

## Data Availability

Data used for analysis is confidential and cannot be shared

## FUNDING

This work was supported by the Inserm cross-cutting program Genomic variability 2018 GOLD.

## Notes

### Competing Interest Statement

The authors have declared no competing interest.

### Author Declarations

We confirm that this study has been declared to INSERM (Institut National de la Santé et de la Recherche Médicale, https://www.inserm.fr/) The data from this study are extracted from the EGB (Echantillon Généraliste de Bénéficiaires), a permanent 1/97 representative sample of the National Health Data System (Système National de Données de Santé, SNDS). The information provided to individuals in EGB on the possible re-use of their data and the procedures for exercising their rights comply with the legislative and regulatory provisions applicable to the processing of personal data in the SNDS. According to French regulation, individuals in SNDS database are informed of the reuse of their data for research and can opposed to this reuse as defined by Articles 92 to 95 of Decree No. 2005-1309 of 20 October 2005 (https://www.legifrance.gouv.fr/loda/article_lc/LEGIARTI000037300884/). As required from French regulation, EGB data can be reuse for research projects from authorized persons once the research project is declared to their institution (INSERM).

