## Supplementary material for "Improving patient clustering by incorporating structured label relationships in similarity measures"

### S1 Choosing thresholds applied in similarity matrices to construct networks

**A.**

| | **Cosine similarity** | **Cosine IDF** | **Cosine WP** | **Cosine Lin** |
|---|---|---|---|---|
| **Threshold** | 0.62403 | 0.59348 | 0.79636 | 0.812705 |
| **Number of female patients** | 5000 | 5037 | 5001 | 5000 |
| **Number of edges** | 77033 | 50827 | 70174 | 57044 |

**B.**

| | **Cosine similarity** | **Cosine IDF** | **Cosine WP** | **Cosine Lin** |
|---|---|---|---|---|
| **Threshold** | 0.66309 | 0.63285 | 0.82437 | 0.833877 |
| **Number of male patients** | 5000 | 5001 | 5000 | 5001 |
| **Number of edges** | 55653 | 39760 | 81077 | 63734 |

Table S1: Choice of the thresholds applied in similarity matrices
Thresholds were identified in the female patient dataset (A) and in the male patient dataset (B)
We computed four similarity matrices, each corresponding to a specific similarity measure. In each matrix, we tested different thresholds ranging from 0.5 to 1 to construct the patient networks. For each threshold tested, we identified the number of patients and edges in the largest connected component of the associated network. To ensure comparable networks, these thresholds were chosen to obtain approximately 5000 patient nodes in the largest connected component of each network. Cosine IDF: Cosine similarity weighted by the Inverse Document Frequency (IDF), Cosine WP: Cosine similarity weighted by the Wu and Palmer measure, Cosine Lin: Cosine similarity weighted by the Lin measure.

[1]These authors contributed equally to this work.

#### S2 Distributions of the four similarity measures computed in the male patient dataset

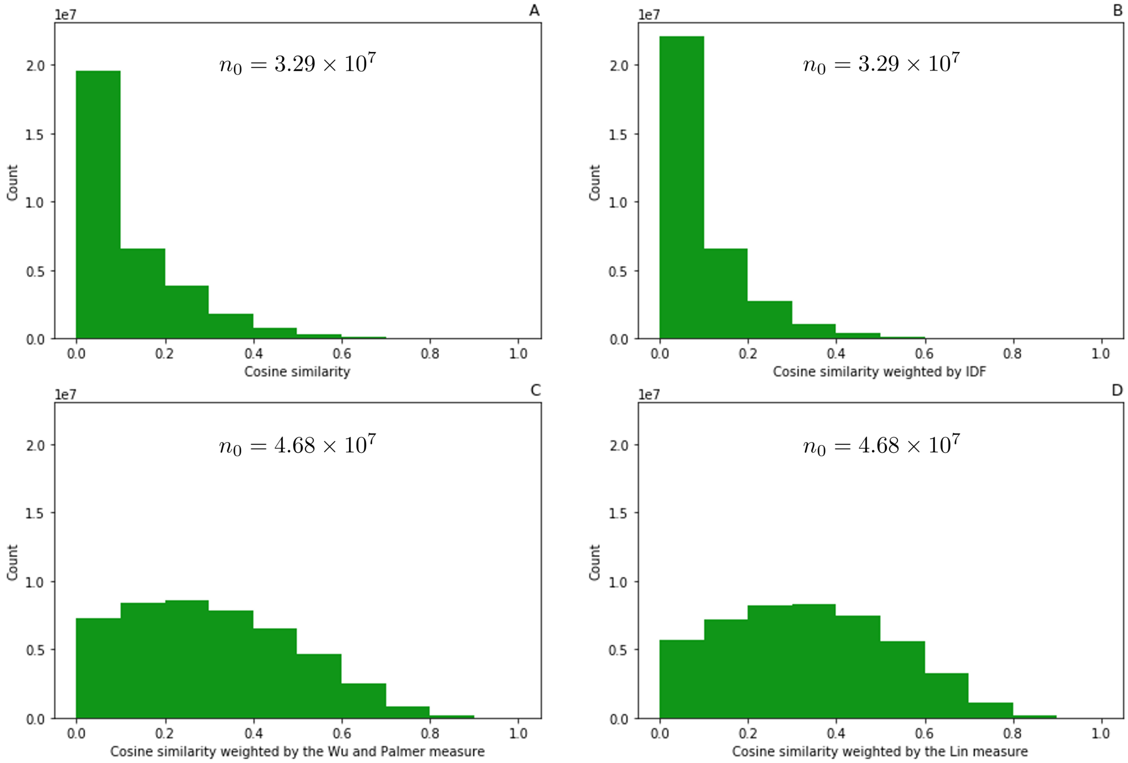


Figure S1: Similarity distributions in the male patient dataset

A: Distribution of the Cosine similarity, B: Distribution of the Cosine similarity weighted by the Inverse Document Frequency (IDF), C: Distribution of the Cosine similarity weighted by the Wu and Palmer measure, D: Distribution of the Cosine similarity weighted by the Lin measure. $n_0$: Total number of pairwise similarities with non-zero values.
